# A high-throughput approach measures cell type-specific telomere lengths in fixed archival tissues from patient cohorts for research on prognosis

**DOI:** 10.1101/2022.01.20.22269597

**Authors:** Christopher M. Heaphy, Reza Zarinshenas, John R. Barber, Christine Davis, Jacqueline A. Brosnan-Cashman, Angelo M. De Marzo, Corinne E. Joshu, Elizabeth A. Platz, Alan K. Meeker

**Affiliations:** Department of Pathology, Johns Hopkins University School of Medicine, Baltimore, MD; Department of Epidemiology, Johns Hopkins Bloomberg School of Public Health, Baltimore, MD; Department of Urology and the James Buchanan Brady Urological Institute, Johns Hopkins University School of Medicine, Baltimore, MD; Sidney Kimmel Comprehensive Cancer Center at Johns Hopkins, Baltimore, MD; Section of Hematology and Medical Oncology, Boston University School of Medicine and Boston Medical Center, Boston, MA; Department of Pathology and Laboratory Medicine, Boston University School of Medicine, Boston, MA

**Author notes:** These authors contributed equally to this study (Christopher M. Heaphy, Reza Zarinshenas). American Association for Cancer Research, Publications Division, Boston, MA.

**Keywords:** Telomeres, fluorescent in situ hybridization, microscopy, archival tissue, prostate cancer, tissue microarray

## Abstract

Telomeres, the repetitive DNA elements at chromosome ends, are pivotal for maintenance of genome integrity. Previous studies from our group and others have highlighted the translational potential of tissue-based telomere length measurements to address the clinical challenge of improving diagnosis, individualized risk stratification, and accurate prognostication of different diseases. Here, we describe a high-throughput method that quantitates cell type-specific telomere lengths at a single cell level in archival tissues from patient cohorts for research on prognosis. This approach is based on telomere-specific fluorescence in situ hybridization (FISH) combined with multiplex immunostaining for cell type-specific antibodies, followed by semi-automated slide scanning and multi-channel acquisition of fluorescent images using the TissueFAXS Plus microscopy workstation and TissueQuest software (TissueGnostics). Here, we demonstrate that this method is sufficiently robust and reproducible to detect biologically significant differences in telomere lengths in archived tissues either on whole slides or sampled across tissue microarrays, which is essential when assessing prognosis in large patient cohorts.

## INTRODUCTION

Telomeres are repetitive DNA (TTAGGG) elements located at the extreme ends of eukaryotic chromosomes and bound by the shelterin protein complex. Telomeres function to mask double strand break DNA damage signals at chromosomal ends and inhibit terminal exonucleolytic degradation, thereby preventing chromosomal fusions (1, 2). Thus, telomeres are critical for maintenance of genome integrity. In normal somatic cells, telomeres shorten with each cell division and significant telomere shortening leads to p53-dependent senescence or apoptosis (3). In contrast, dysfunctional telomeres and the abrogation of cell cycle checkpoints during malignant transformation allows genomic instability to ensue via chromosomal breakage-fusion-bridge cycles (4).

Since accumulation of genomic instability promotes tumorigenesis and progression, numerous investigators have utilized tissue-based telomere length measurements to evaluate cancer prognosis, as well as risk, across a variety of tumor types [reviewed in (5, 6)]. For example, our group previously demonstrated that telomere length and cell-to-cell variability in telomere lengths are useful in predicting prostate cancer death in men surgically treated for their clinically localized prostate cancer (7). We have also shown that stromal cell telomere shortening is associated with an increased risk of prostate cancer (8). Taken together, these findings highlight the translational potential of tissue-based telomere measurements for cancer prognostication and risk stratification.

Here, we validate a combined telomere-specific FISH and multiplex immunofluorescence staining method, followed by semi-automated slide scanning and multi-channel acquisition of digitized fluorescent images, to measure cell type-specific telomere lengths in archival human tissues from patient cohorts for research on prognosis.

## MATERIALS AND METHODS

### Telomere-specific FISH and Immunostaining

The staining protocol is performed similar to previous descriptions (9, 10), with the following modifications (see **Supplemental Methods**). Briefly, a 4 µm formalin-fixed, paraffin-embedded tissue section is deparaffinized, hydrated, and steamed in target retrieval citrate buffer. Telomere-specific peptide nucleic acid (PNA) probe (CCCTAACCCTAACCCTAA with the N-terminal covalently linked to Cy3) is applied, denatured by incubation for 5 min at 84°C, and hybridized overnight. For cell type-specific identification, immunofluorescent staining is then performed. After appropriate washing, the nuclei are 4’-6-diamidino-2-phenylindole (DAPI) stained, slides are mounted with Prolong anti-fade, and coverslipped.

### Microscopy and Image Analysis

For automated scanning and image acquisition, the TissueFAXS Plus 6.108 automated microscopy workstation (TissueGnostics) is utilized, which contains an 8-slide ultra-precise motorized stage and utilizes a Zeiss Z2 Axioimager microscope. For the image analysis, a separate high-performance workstation with TissueQuest 6.120 software module (TissueGnostics) is used to analyze the digitized fluorescent images with precise nuclear segmentation.

## RESULTS

### Development of a combined telomere-specific FISH and multiplex immunofluorescence staining method

In combination with telomere-specific FISH, multiplex immunostaining for cell type-specific antibodies is performed to identify specific cell types. For example, in benign and cancerous prostate tissue as shown in **Figure 1**, a basal epithelial cell-specific anti-cytokeratin primary antibody (i.e. CK903) is used to identify the benign prostate glands by delineating the basal prostate epithelial cells from the luminal prostate epithelial cells. Simultaneously, to easily identify and distinguish normal luminal epithelial cells and prostate cancer cells from the surrounding stromal cells, anti-NKX3.1 and/or anti-FOXA1 primary antibodies are included. Additionally, to identify and easily exclude lymphocytes in tissue, anti-CD3 and anti-CD20 primary antibodies are used.

**Figure 1.**
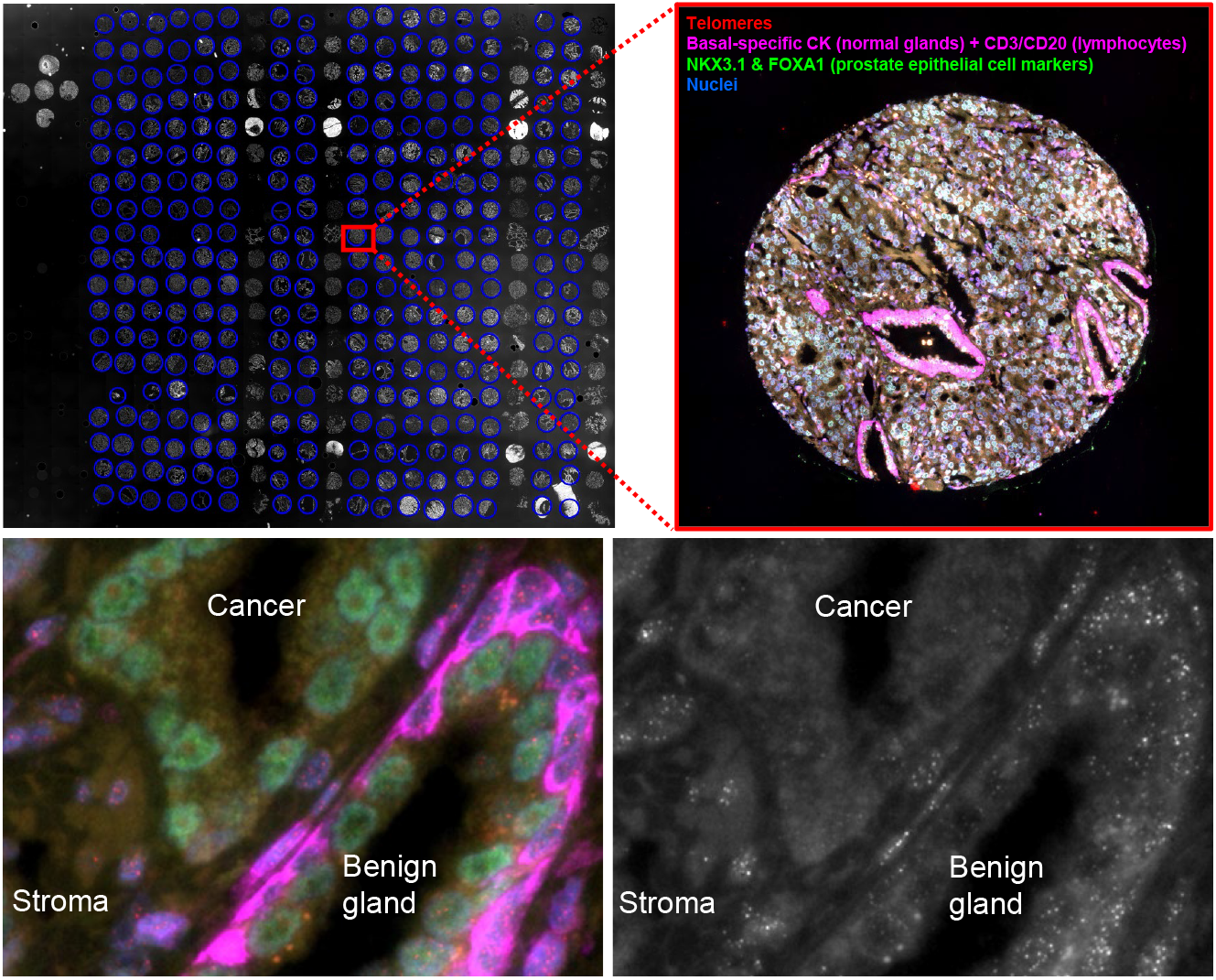
Representative example of automated scan and image acquisition of a stained tissue microarray. The upper left panel shows a 10X preview image captured with the DAPI channel to allow for accurate orientation. The upper right panel highlights a specific region of interest (e.g. tissue microarray spot) in which multiple sequential images are captured with a 40X oil objective using the DAPI, GFP, Cy3, and Cy5 channels. The bottom panels highlight a representative region containing cancer and benign glands (multicolor combined immunofluorescence plus telomere FISH, left) and highlighting dramatically reduced telomeric FISH signals in the cancer cells compared to the benign cell populations (greyscale image of the Cy3 telomere FISH channel, right).

### Measurement of telomere length in individual cells of specified type

Using the automated microscopy workstation, a 10X preview image on DAPI is first captured to allow for accurate orientation (**Figure 1**). Next, the regions of interest are identified and images are captured with a 40X oil objective using, sequentially, the DAPI, GFP, Cy3, and Cy5 channels (**Figure 1**). To optimally acquire the telomere signals, the extended focus parameter is used with 3 steps above and below the z-axis plane, with a step size of 0.8 microns for each step. Using this approach, an entire TMA that contains ∼400 spots can be imaged overnight (∼14 hours), which is considerably faster than other current telomere-based imaging modalities (11).

For the image analysis, as shown in **Figure 2**, a separate high-performance workstation is used to analyze the digitized fluorescent images with precise nuclear segmentation. Annotated regions of interest are set and processed. If required, based on the antibody staining, any necessary exclusion areas can be excluded from the analysis. An exclusion region can be drawn either before or after an analysis has been performed. Relative telomere lengths are determined by calculating the ratio of the total Cy3 telomere FISH signal intensity to the total DAPI intensity for each nucleus, thereby compensating for differences in nuclear cutting planes and ploidy.

**Figure 2.**
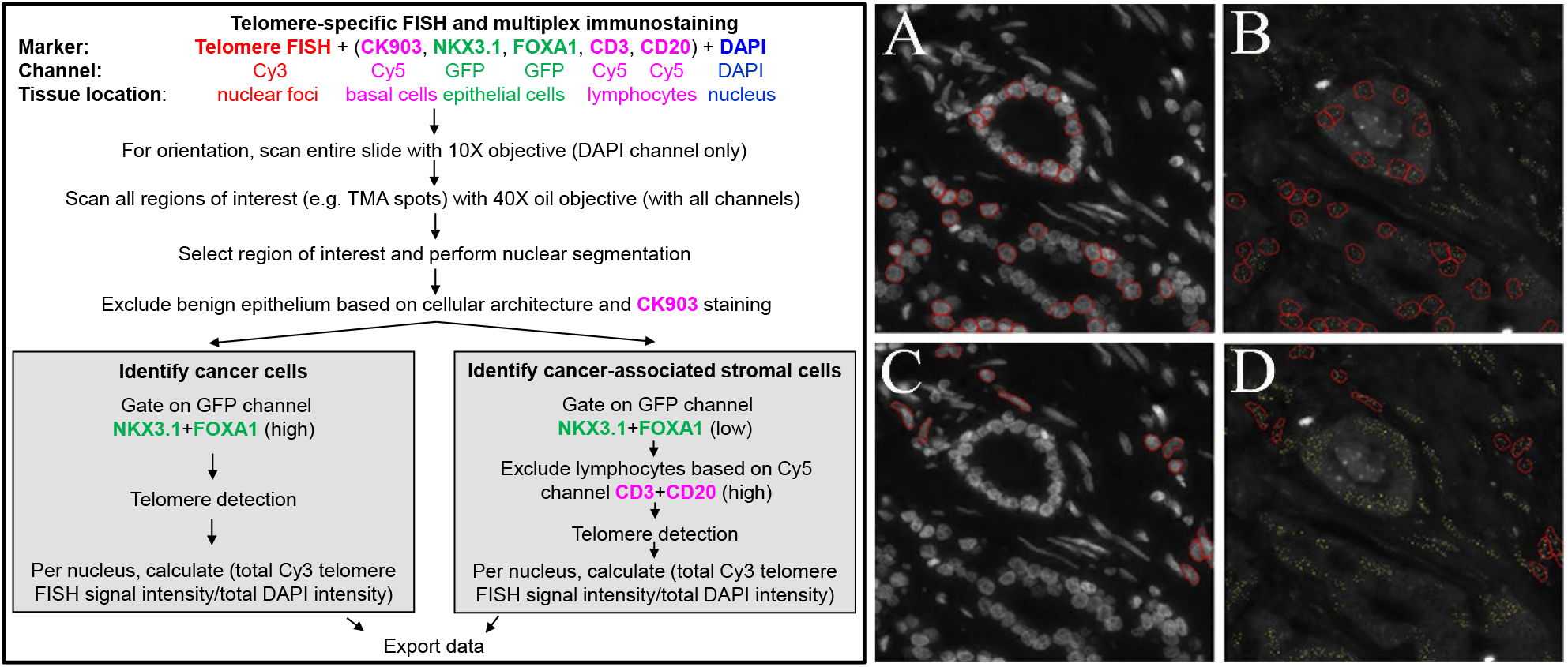
Overall workflow of the method and example of automated prostate cancer cell and cancer-associated stromal cell segmentation and telomere detection. On the left, the overall workflow of staining, scanning, nuclear segmentation, cell-type identification, and telomere detection is depicted. On the right panel, after exclusion of benign prostate glands based on CK903 staining and cellular architecture, a representative example of the nuclear segmentation parameters that specifically identify (A, B) cancer cell nuclei (red circles; upper panels) based on high GFP or (C, D) cancer-associated stromal cell nuclei (red circles; lower panels) based on low GFP and Cy5 (i.e. lymphocytes) exclusion. A specialized algorithm to detect telomere FISH signals (yellow dots; right panels) is shown. Relative telomere lengths are determined by calculating the ratio of the total Cy3 telomere FISH signal intensity to the total DAPI intensity for each nucleus.

### Telomere-specific FISH signal intensities are linearly proportional to telomere length and can be quantified via digital image analysis

In order to validate the method to detect differences in telomere lengths, a HeLa cell line bearing a single copy of the dox-inducible TPP1 gene (i.e. HeLa TPP1) that causes telomere elongation when expressed (12) were continually grown in the presence of dox, and collected at different time points (populations doublings: 0, 28, 58, and 84), thereby possessing different telomere lengths. A standard formalin-fixed, paraffin-embedded cell block containing pellets from each time point was created, stained, imaged, and analyzed using this new method. Cells from each corresponding time point were also collected and analyzed independently by telomere length by Terminal Restriction Fragment (TRF) Southern blot analysis, the “gold-standard” method for determining absolute telomere lengths (13). As shown in **Figure 3**, the telomere length ratios determined by digital image analysis corresponded linearly with absolute telomere lengths as determined by TRF analysis (R^2^=0.98). Thus, these data also allow for telomere length ratios to be extrapolated to absolute telomere lengths.

**Figure 3.**
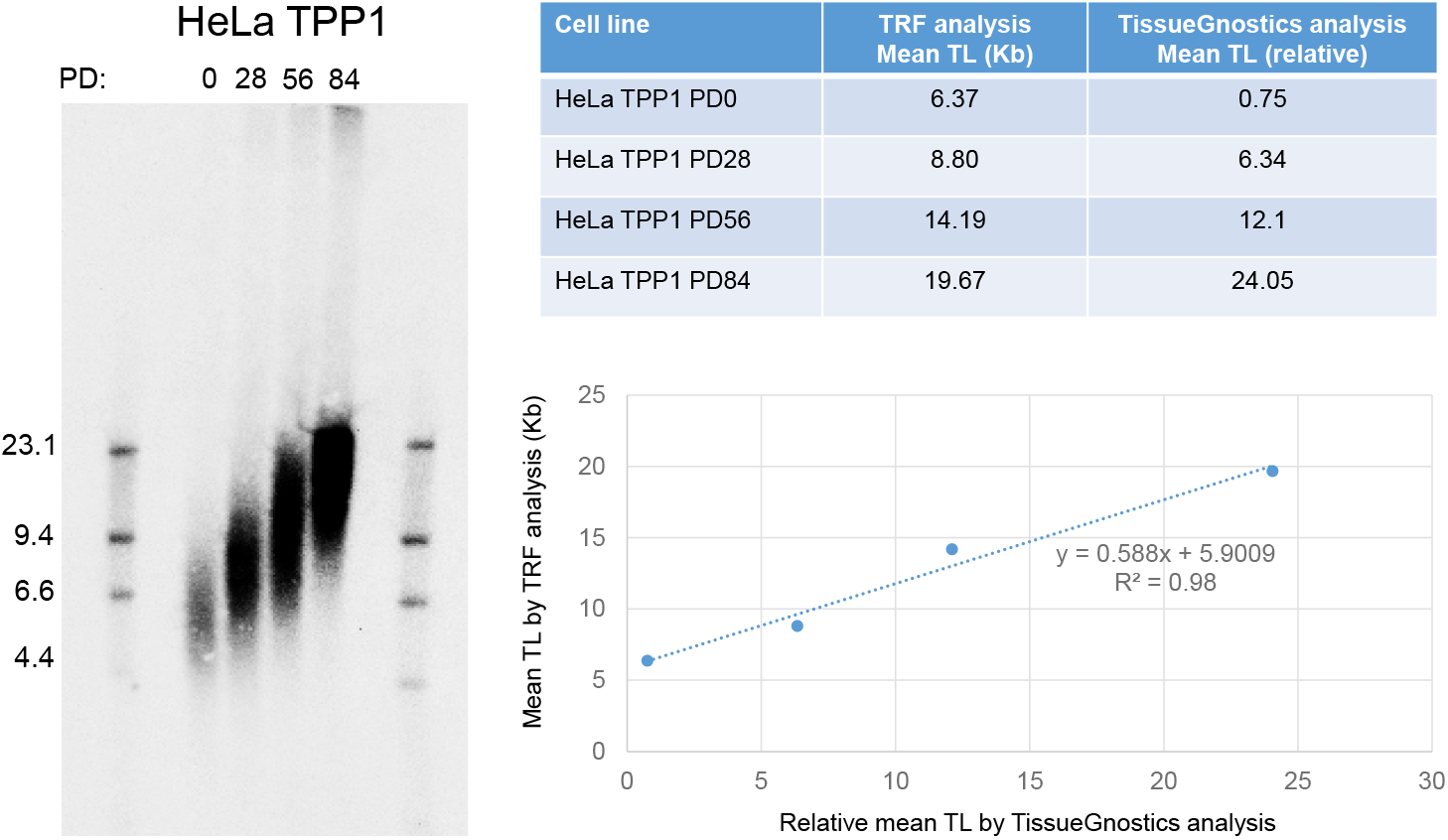
Terminal Restriction Fragment (TRF) Southern blot analyses reveals a wide range of telomere lengths in the HeLa TPP1 cell line, increasing over time with population doubling when grown in the presence of dox. The telomere length (TL) ratios determined by digital image analysis on FISH stained slides using the TissueGnostics approach corresponds linearly with the TRF Southern blot analyses.

## DISCUSSION

Members of our group previously developed the TELI-FISH method which allows for telomere length determination in individual cells (9). However, this method is not automated and requires substantial labor for image collection and image analysis, thereby limiting the utility of further testing and application of tissue-based telomere measurements in a clinical setting. In contrast, other methodologies such as Terminal Restriction Fragment (TRF) Southern blot analysis (13, 14), telomere-specific qPCR (15, 16), Single Telomere Length Analysis (STELA) (17), Telomere Shortest Length Assay (TeSLA) (18), or estimating telomere lengths from whole genome sequencing data (e.g. TelSeq) (19) requires less labor, although cellular identities and histologic architecture of the tissue, and spatial resolution are completely lost (20).

Our new approach is based on telomere-specific FISH combined with cell type-specific immunofluorescent staining, followed by semi-automated slide scanning and multi-channel acquisition of digitized fluorescent images using the TissueFAXS Plus microscopy workstation, followed by image analysis using TissueQuest software (TissueGnostics). This new method is sufficiently robust and reproducible to detect biologically significant differences in telomere lengths in archived human tissues.

## Data Availability

All data produced in the present study are available upon reasonable request to the authors.

## ACKNOWLEDGMENTS

This research was supported by grants from the Department of Defense Prostate Cancer Research Program (W81XWH-12-1-0545), the National Cancer Institute (P50 CA058236, P30 CA006973), and the Prostate Cancer Foundation (Young Investigator Awards to C. Joshu, C. Heaphy). The content is solely the responsibility of the authors and does not necessarily represent the official views of the National Institutes of Health.

