## Supplemental Methods for "A high-throughput approach measures cell type-specific telomere lengths in fixed archival tissues from patient cohorts for research on prognosis"

#### Reagents and Materials

- Formalin-fixed paraffin-embedded clinical tissue specimens (4 µm thick sections cut onto charged slides)
- VistaVision HistoBond (VWR; Cat #16004-406) or other positively charged slides
- Xylene
- Graded ethanol series (70%, 95%, 100%)
- Antigen unmasking solution (Vector Laboratories; Cat #H-3300)
- Hybridization probe mix (see recipe)
- Telomere-specific peptide nucleic acid (PNA) probe: CCCTAACCCTAACCCTAA with the N-terminus covalently linked to Cy3 (Panagene; Cat #F1002)
- Pan-centromere-specific PNA probe with the N-terminal covalently linked to Alexa Fluor 488 (Panagene; Cat #F3012)
- Formamide (Sigma; Cat # F-7503)
- 1M Tris pH 7.5 (Quality Biological; Cat #351-006-101)
- Coverslips (Fisher; Cat #12-545-B)
- Humidified chamber for hybridization
- Wash buffer (see recipe)
- Phosphate buffered saline with Tween 20 (PBST); (Sigma; Cat #P-3563)
- Serum-free protein block (DAKO; Cat #X0909)
- Antibody dilution buffer (Ventana; Cat #ABD250)
- Basal-specific anti-cytokeratin primary antibody (34BE12, Enzo; Cat #ENZ-C34903)
- Anti-NKX3.1 primary antibody (Athena; Cat #0314)
- Anti-FOXA1 primary antibody (Abcam; Cat #ab23738)
- Anti-CD3 primary antibody (clone F7.2.38, DAKO; Cat #M7254)
- Anti-CD20 primary antibody (Abcam; Cat #ab9475)
- Phosphate buffered saline (PBS); (Quality Biological; Cat #114-058-101)
- Anti-rabbit IgG fraction Alexa Fluor 488 (Thermo Fisher Scientific; Cat #A21441)
- Anti-mouse IgG fraction Alexa Fluor 647 (Thermo Fisher Scientific; Cat #A21237)
- 4′-6-diamidino-2-phenylindole (DAPI); (Sigma; Cat #D-8417)
- Prolong anti-fade mounting media (Life Technologies; Cat #P36934)
- Zeiss Immersol 518F Immersion Oil (Fisher Scientific; Cat #12-624-66A)

**PART I – Telomere-specific FISH and Immunolabeling**

This part of the protocol describes the use of telomere-specific fluorescence *in situ* hybridization (FISH) combined with immunolabeling to identify specific cell populations within archived formalin-fixed paraffin-embedded (FFPE) tissue specimens. This protocol provides single cell resolution of telomere length in specific cell-types, while preserving tissue architecture. To highlight the potential clinical utility of the assay, the protocol outlines an example the specific use of telomere-specific FISH combined with immunostaining for prostate basal epithelial cell-specific cytokeratin, prostate luminal epithelial-cell specific nuclear markers (NKX3.1 and FOXA1), and lymphocyte-specific markers (CD3 and CD20) in benign and cancerous human prostate tissue. However, as outlined below, validated antibody combinations can be readily optimized for other applications in a variety of benign or diseased tissue types (e.g. breast, pancreas, brain, etc.) specific for particular cell types of interest for inclusion and/or exclusion. Different variations of this part of the protocol have been previously used for qualitative and quantitative telomere length assessment; thus, many of these staining steps have been previously described [1-3].

1. Obtain standard unstained tissue sections (4 μm) from tissue type of interest (or tissue microarrays; TMAs) from FFPE tissue blocks on positively charged slides.
   ***Hint****: This protocol can be used on any fixed tissue specimen – surgical resection, biopsy, or constructed TMAs. Additionally, a standard slide that contains specimens/cells with known telomere lengths can be included in the analysis to allow for direct conversion of measured telomere ratios in tissue samples to actual average telomeric DNA lengths.*
2. Place slides on a pre-heated 65°C slide warmer for 10 min.
3. Wash slides three times (5 min/wash) in xylene, followed by twice (3 min/wash) in 100% ethanol, twice (3 min/wash) in 95% ethanol, once in 70% ethanol (3 min), once in deionized distilled water ($ddH_20$; 3 min), and once in 1% Tween 20 (1 min).
   ***Attention****: These washes, and all washes described below unless specifically stated, should be completed with gentle agitation at room temperature.*
4. Incubate slides in a plastic slide holder with pre-heated antigen unmasking solution in a pre-heated (boiling) kitchen steamer for 30 min.
5. Remove container from the steamer, carefully open the lid of the container, and let cool at room temperature for 10 min.
6. Wash slides once (1 min) in $ddH_20$, twice (3 min/wash) in 70% ethanol, twice (3 min/wash) in 95% ethanol, and once (2 min) in 100% ethanol. Allow the slides to completely air dry.
7. Add 35 μl of probe mix (0.33 μg/ml Cy3-conjugated telomere-specific PNA probe) to each slide.
   ***Attention****: If an additional positive hybridization control is desired, a pan-centromere-specific PNA probe can also be included in the probe mix at the same concentration as the telomere PNA probe.*

8. Coverslip each slide and place slides at 84°C on a pre-heated temperature-controlled heat block for 5 min.
   ***Attention**: Be careful to avoid the presence of air bubbles when placing the coverslip. Additionally, check to ensure that the entire tissue specimen is covered by the probe mix.*
9. Hybridize slides for at least 2 hr at room temperature in a dark, humidified chamber.
   ***Hint**: If desired, slides can also be hybridized overnight at room temperature in a dark, humidified chamber.*
10. After hybridization, gently remove the coverslip from each slide with forceps and wash the slides twice (15 min/wash) in wash buffer (see recipe).
11. Wash slides three times (5 min/wash) in PBST.
12. Apply 2-3 drops (~200 µl) of serum-free protein block onto each slide and incubate in a humidified chamber at room temperature for 30 min.
13. Wash slides twice (5 min/wash) in PBST.
14. For cell type-specific identification, appropriately dilute primary antibodies of interest in antibody dilution buffer.
    As a specific example, to accurately identify basal cells, epithelial cells, and lymphocytes in benign and cancerous prostate tissue, use the following diluted primary antibodies:
    a. basal-specific anti-cytokeratin primary antibody (1:50 dilution)
    b. anti-NKX3.1 primary antibody (1:1000 dilution)
    c. anti-FOXA1 primary antibody (1:500 dilution)
    d. anti-CD3 primary antibody (1:200 dilution)
    e. anti-CD20 primary antibody (1:20 dilution)
    ***Hint**: Replacing these specific antibodies with other validated antibodies can be performed to selectively identify cells of particular type for inclusion or exclusion in the image analysis determination of telomere length.*
15. Apply 200 µl of diluted primary antibody mix onto each slide and incubate at room temperature for 2 hrs in a humidified chamber.
16. Wash slides three times (5 min/wash) in PBST.
17. Based on the primary antibodies used in step 14, dilute (1:100) the fluorescent secondary antibodies together in PBS.
    As a specific example, to accurately detect the antibody mix (outlined in step 14) in benign and cancerous prostate tissue, use the following diluted secondary antibodies:
    a. anti-rabbit IgG fraction Alexa Fluor 488
    b. anti-mouse IgG fraction Alexa Fluor 647
    ***Hint**: Deciding which secondary antibodies to use (e.g. species or fluorescent labels) is dependent on the primary antibodies chosen and the localization patterns of each protein of*

*interest in the tissue type of interest (i.e. nuclear, cytoplasmic, membrane).*

18. Apply 200 µl of diluted secondary antibody mix onto each slide and incubate at room temperature for 30 min in a humidified chamber.
19. Wash slides three times (5 min/wash) in PBST and once (5 min) in $ddH_20$.
20. Incubate slides at room temperature in 4′-6-diamidino-2-phenylindole (DAPI) solution (500 ng/ml in $ddH_20$) for 10 min.
    ***Attention****: DAPI staining needs to be precise in terms of temperature of the solution and incubation time. To accomplish this, all slides in an experiment can be placed in a slide holder and submerged in a plastic container holding 200 mls of room temperature DAPI solution. The entire container can then be incubated with gentle agitation for 10 min at room temperature, thereby allowing for consistent DAPI staining from slide to slide.*
21. Wash slides three times (5 min/wash) in $ddH_20$ and then drain excess water for each slide.
    ***Attention****: While removal of excess water is necessary, do not allow the slides to completely dry during this step.*
22. Apply 2-3 drops (~40 µl) of Prolong antifade mounting medium to each slide and carefully apply a coverslip to each slide.
    ***Attention****: Be careful to avoid air bubbles when placing the coverslip. If an air bubble is present over the tissue area after the coverslip is applied, do not try to “pop” the bubble by pressing on top of the coverslip. Simply, remove the coverslip gently with forceps, and add additional Prolong and repeat the coverslip process.*
23. Store slides at room temperature (in the dark) for 24 hr to allow the Prolong to harden and then store the slides at 4°C for at least 1 week to allow the Prolong to completely cure, prior to proceeding to PART II.

**PART II – Microscopy using TissueFAXS**

This part of the protocol describes how individually stained tissue slides, in particular individual TMA spots on the TMA slides, are imaged, and digital images captured, using the TissueFAXS Plus (Tissue Gnostics) automated microscopy workstation and Zeiss Z2 Axioimager microscope.

1. Turn on the computer, microscope (Imager.Z2; Zeiss), and X-Cite Series 120 PC lamp.
   ***Hint****: In general, another brand of fluorescence microscope can be substituted, although the exact exposure times may vary and the image capture process may differ than as described below.*
2. Open the TissueFAXS program, perform the calibration when prompted, and create a new fluorescent experiment with the following parameters:
   a. Preview Objective = 10X, Air
   b. Acquisition Objective = 40X, Oil

c. Select the DAPI, GFP, CY5, CY3 filters/reflectors
d. Confirm camera is PCO PixelFly / 0

***Attention**: Telomere detection requires the use of at 40X objective or 63X objective for acquisition. While a 10X objective is used to generate the low-resolution preview image, the telomeres cannot be readily distinguished in human tissue using a 10X or 20X objective.*

3. Place slide(s) snugly into stage, select the DAPI reflector, and adjust the exposure time accordingly for an optimal 10X low resolution preview scan.
   ***Hint**: A reasonable exposure time for the preview scan is 80 ms (for DAPI) with the lower and upper sensitivity thresholds set to [500, 4095] at 50% lamp power.*
4. Next, apply immersion oil to each slide and select the 40X objective and DAPI reflector as the focus channel.
5. Adjust exposure times accordingly for each reflector, while avoiding saturation of single intensities for the DAPI and CY3 reflectors, in particular.
   As a specific example, to accurately identify the telomeres in basal cells, epithelial cells, and lymphocytes in benign and cancerous prostate tissue, use the following exposure times and lower and upper sensitivity thresholds at 50% lamp power (X-Cite 120PC Q Illuminator):
   DAPI = 3ms [300, 4095]
   GFP = 70ms [800, 4095]
   CY5 = 200ms [500, 4095]
   CY3 = 140ms [300, 4095]
6. For each slide, add any regions of interest or set the appropriate TMA spot configuration.
   To optimally acquire the telomere signals, utilize the extended focus parameter and the number of steps to 3 (above and below) and step size to 0.8 microns.
7. Finally, select “Acquire”. When all images have been acquired, save the project accordingly.

**PART III – Telomere Analysis using TissueQuest**

This part of the protocol describes how the digitized fluorescent telomere FISH signals in the saved image files are quantified using the TissueQuest software. Relative telomere lengths in individual cells can be determined by calculating the ratio of the total CY3 intensity by the total DAPI intensity for each identified nucleus.

1. Import a TissueFaxs project and save the new project in the desired location. Add “Dots Virtual Marker” for subsequent telomere detection.
   ***Hint**: In general, another software that can perform nuclear segmentation and thoroughly analyze digitized fluorescent images can be substituted, although the exact process may vary.*

2. Choose a region of interest and add new regions of interest (ROI) through the free draw function.
3. Based on the antibody staining, set any necessary areas to be excluded from the analysis. In the specific example of benign and cancerous prostate tissue, the basal-specific anti-cytokeratin staining is used to identify normal prostate glands. In contrast, the NKX3.1 and FOXA1 nuclear staining identifies cancer cells for benign epithelial cells. Finally, CD3 and CD20 staining identifies the presence of lymphocytes.
   ***Hint****: Exclusion regions can be drawn either before or after an analysis has been performed.*
4. For the analysis, adjust the parameters for each of the DAPI, GFP, CY5, and CY3 reflectors accordingly.
   To analyze a specific ROI, select "Analyze". To view the results of nuclear segmentation, select the DAPI tab and "Shades Overlay".
5. Creating a Scattergram:
   a. A new scattergram of interest can be added through "Manage Diagrams".
   b. Choose desired markers and parameters for the X-axis and Y-axis.
   c. Choose desired Input Gates and specific cut-off values.
   d. Gates can be linked consecutively to identify cells of interest.
   
   ***Hint****: The scattergram function and linking gates together is analogous to preforming flow cytometry in fixed tissue specimens.*
6. Viewing the raw data corresponding to an analyzed region can be displayed by viewing the backward data in a specific scattergram.
   a. Each row represents data from an individually identified nucleus.
   b. CY3 dot sum intensity and DAPI sum intensity data (or another parameter of interest) can be added to the column header.
7. Export all data to Excel spreadsheets and convert these spreadsheets into a format suitable for statistical analysis, such as a SAS dataset.
8. For each individual and for each cell, calculate the ratio of CY3 dot sum intensity and DAPI sum intensity and multiply by 1000; this ratio is the telomere ratio (TR).
9. For use in any precision medicine application, the median or mean of TR for an individual may be used. In addition, the distribution of TR among an individual's cells of a specific type can be expressed using measures of central tendency, spread, shape, proportion under or over a cutpoint or set of cutpoints. Additionally, these measures of the TR distribution in any given cell type or in various combinations with other cell types can be used, for example, to predict risk of disease or death from a disease or health condition in those without the diagnosis, risk of poor outcome (e.g., progression, death) in those with the diagnosis, and overall survival.

### Recipes

Probe mix

0.33 µg/mL of Cy3-conjugated telomere-specific PNA probe

0.33 µg/mL of Alexa Fluor 488-conjugated pan-centromere-specific PNA (if desired)

70% formamide

10 mmol/L Tris, pH 7.5

$ddH_20$

Wash Buffer

70 ml formamide

29 ml $ddH_20$

1 ml 1M Tris, pH=7.5

### Equipment

- Slide warmer with temperature control
- Kitchen steamer
- ThermoBrite denaturation and hybridization system (VWR)
- Z2 Axioimager microscope (Zeiss)
- X-Cite Series 120 fluorescence illumination microscope light source (Lumen Dynamics)
- PixelFly high performance digital CCD camera (PCO)
- TissueFAXS Plus 6.108 automated microscopy workstation (Tissue Gnostics)
- TissueQuest 6.120 software module (Tissue Gnostics)
